# Traumatic Brain Injury Increases Risk of Psychiatric Disorders Following Neurological Disease: Evidence from the UK Biobank Cohort

**DOI:** 10.64898/2026.07.02.26357128

**Authors:** Grace Revill, Norman Poole, Christina Carlisi, Anthony S. David, Vaughan Bell

## Abstract

**Background:** Traumatic brain injury (TBI) is common in older adults and frequently co-occurs with neurological conditions such as dementia, stroke, and Parkinson’s disease. Both TBI and neurological disease independently increase vulnerability to psychiatric disorders, yet the long-term impact of lifetime TBI exposure on mental health outcomes within these neurological populations remains poorly understood.

**Methods:** Using data from the UK Biobank cohort (N = 502,154), we examined associations between TBI and subsequent psychiatric diagnoses in individuals diagnosed with dementia, stroke, or Parkinson’s disease. Participants were assigned to one of four exposure groups (no TBI and no neurological disease, TBI only, neurological disease only, TBI plus neurological disease), and mixed-effects logistic regression estimated psychiatric risk across these groups and within each neurological condition. Kaplan-Meier curves and Cox proportional hazards models assessed 10-year cumulative risk. All analyses were adjusted for demographic and socioeconomic factors.

**Results:** The combined presence of TBI and neurological disease produced the greatest psychiatric risk, exceeding that observed for either condition alone. Across dementia, stroke, and Parkinson’s disease, TBI most reliably increased the odds of mood disorders, while associations for anxiety were less consistent. Ten-year cumulative incidence analyses demonstrated higher risk of mood and overall mental health disorders across all three neurological cohorts.

**Conclusions:** Lifetime TBI exposure is associated with increased likelihood and long-term risk of psychiatric disorders in individuals with dementia, stroke, and Parkinson’s disease, with mood disorders being the most consistent outcome across conditions. These findings highlight the importance of recognising TBI history when assessing and managing psychiatric risk in ageing neurological populations and underscore the need for integrated clinical pathways that address both neurological and mental health needs.

## Introduction

Traumatic brain injury (TBI) is a major public health concern and one of the leading causes of disability worldwide, affecting millions of individuals across the lifespan (Maas et al., 2022). Although TBI occurs at all ages, older adults now represent the group with the highest rates of TBI-related hospitalisation and mortality, largely due to age-related frailty and increased risk of falls (Gardner et al., 2018; Maas et al., 2022). Clinical management in this population is particularly complex, as TBI frequently co-occurs with chronic neurological and physical conditions, which can mask symptoms, complicate diagnosis, and heighten vulnerability to poor outcomes (Dams-O’Connor et al., 2020; Gardner et al., 2018). Despite this growing burden, older adults with TBI remain comparatively underrepresented in research.

TBI comprises a wide spectrum of severity, from mild to severe, and can lead to outcomes ranging from transient symptoms to persistent, long-term impairments (Bramlett & Dietrich, 2015; Wilson et al., 2017). TBI is strongly associated with a spectrum of psychiatric sequelae, including depression, anxiety and psychotic disorders (Howlett et al., 2022). These psychological consequences can persist long after the initial injury, impacting daily functioning, quality of life, and long-term burden on healthcare systems (Howlett et al., 2022; Kureshi et al., 2023).

Psychiatric conditions such as depression, anxiety, and psychotic disorders are also common in individuals with chronic neurological conditions, including dementia (Orgeta et al., 2022), stroke (Zhang et al., 2020), and Parkinson’s disease (Gallagher & Schrag, 2012). In these populations, psychiatric comorbidities are linked to poorer clinical outcomes and accelerated disease progression (Mo et al., 2023; Zhou et al., 2023), higher levels of caregiver strain (Geerlings et al., 2023; Pucciarelli et al., 2017), and reduced overall quality of life (Burchill et al., 2024; Prisnie et al., 2018).

Notably, TBI itself is an established risk factor for the later development of Parkinson’s disease (Gardner et al., 2015), stroke (Karamian et al., 2025) and dementia (Simmonds et al., 2025). However, the role of TBI as a risk factor for psychiatric complications within these other common neurological disorders is poorly understood. It therefore remains unclear whether comorbid TBI modifies the clinical phenotype of these disorders by increasing psychiatric symptom burden, or whether it merely elevates the likelihood of their occurrence without substantively altering the presentation. These issues are particularly relevant in older adults, who experience the highest rates of TBI-related hospitalisation, frequently from preventable falls (Cusimano et al., 2020), and carry the greatest burden of neuropsychiatric comorbidities in neurological disorder (Berrechid, 2023; Maas et al., 2022).

One obstacle in understanding these issues has been the scarcity of large-scale, population-based datasets large enough to address the combined impact of TBI and neurological disease on mental health outcomes. Many existing cohorts are underpowered, particularly when examining multiple neurological conditions and psychiatric outcomes simultaneously, and short follow-up periods further constrain the ability to capture long-term consequences of lifetime TBI exposure. To overcome these limitations, large longitudinal studies are needed to characterise the long-term consequences of lifetime TBI exposure across multiple neurological conditions and psychiatric outcomes across diverse patient populations.

The UK Biobank dataset (Allen et al., 2024) offers a unique opportunity to address these gaps. Its extensive cohort of over 500,000 middle-aged and older adults, provides detailed information on lifetime TBI exposure, comprehensive mental health assessments, and linkage to hospital and death records (Allen et al., 2024; Sudlow et al., 2015). This population allows for the investigation of how TBI influences psychiatric outcomes during a life stage when neurological conditions commonly emerge and when long-term consequences are most likely to manifest. The large sample size further enables simultaneous analysis across multiple neurological conditions while controlling for potential confounding factors, enhancing the generalisability and robustness of findings.

Consequently, this study aimed to understand to what extent TBI is a risk factor for the development of subsequent neuropsychiatric complications in dementia, stroke and Parkinson’s disease using UK Biobank cohort data.

## Methods

### Dataset

UK Biobank (UKB) is a large, prospective cohort of over 500 000 generally healthy adults aged 40–70 years (mean = 56.5, SD = 8.1) at recruitment (2006–2010), registered with the UK National Health Service and assessed at one of 22 recruitment centres (Allen et al., 2024). Details of participant recruitment for UKB have been described previously (Sudlow et al., 2015). Written informed consent was obtained from all participants prior to study initiation, with access to UK Biobank data approved by the UK Biobank Access Committee (Application number 117100).

### Participants

All-cause dementia, stroke, and Parkinson’s disease were identified using algorithmically defined outcomes derived from linked hospital admissions, death registries, and participant self-reported medical information, as previously described and validated elsewhere (Allen et al., 2024; Rannikmäe et al., 2020; Wilkinson et al., 2019). These algorithms integrate data from multiple sources to establish the date of diagnosis. For this study, three distinct cohorts were constructed, each comprising individuals diagnosed with dementia, stroke, or Parkinson’s disease, respectively.

### Outcomes and exposures

#### TBI

History of TBI was captured by two means: (i) via self-reported ‘head injury’ at baseline (2006-2010); and (ii) via capture of data on 53 ICD codes specific to TBI and associated terms (Supplementary Table 1). Primary and secondary ICD-10 diagnoses were based on hospital episode and admission (HES) admission NHS data, as described in the open-access protocol (https://www.ukbiobank.ac.uk/wp-content/uploads/2025/01/Main-study-protocol.pdf), from May 1995 to October 2022. The date of injury was defined primarily as the earliest date of the 53 ICD codes specific to TBI. Following Lyall et al. (2024), for participants with ICD codes but no specific date, the earliest inpatient diagnosis date was used as a proxy inpatient date.

#### Psychiatric disorders

Mood, anxiety, and psychotic disorders were identified using ICD-10 diagnostic codes, drawing on data from inpatient hospital records, participant self-reported questionnaires, and death registry records, as previously described elsewhere (Davis et al., 2019; Li et al., 2022; Stroganov et al., 2022). The mood disorder category included depressive episode, recurrent depressive disorder, bipolar disorder, affective disorder, or other specified mood/affective disorder. Anxiety disorders included any diagnosis of phobic disorder, obsessive-compulsive disorder (OCD), or other specified anxiety disorders. Psychotic disorders comprised schizophrenia, schizoaffective disorder, delusional disorder, or acute or transient psychotic disorder. In addition, a composite category, ‘any mental health disorder,’ was defined as the presence of any mood, anxiety, or psychotic disorder.

### Covariates

Self-reported sex, age, race and ethnicity, socioeconomic status (Townsend Deprivation Index) and alcohol intake frequency at baseline were included as covariates in adjusted analyses as they have all been shown to be independent predictors of TBI and psychiatric problems (Corrigan et al., 2025; Kirkbride et al., 2024; Miller et al., 2021; Puddephatt et al., 2022; Weil et al., 2018). The 5-level race and ethnicity variable (‘Asian or Asian British’, ‘Black, Black British, Caribbean or African’, ‘Mixed or Multiple Ethnic Groups’, ‘White’, and ‘Other’) was constructed based on the ethnic group classification used in the 2021 Census (*Ethnic Group, England and Wales - Office for National Statistics*, n.d.). The Townsend Deprivation Index is derived from participants’ postal codes at recruitment, combining four census-based indicators: unemployment, non-car ownership, non-home ownership, and household overcrowding. Higher scores indicate greater material deprivation (Ye et al., 2021).

### Analysis

For the purposes of this study, TBI exposure was defined using harmonised self-reported and ICD-coded information. To ensure appropriate temporal ordering, only ICD-coded TBI that clearly preceded the onset of mental health conditions were retained. Participants were required to have at least one ICD TBI code and a valid injury date, and were kept if they met one of the following criteria: (i) they had no recorded mental health diagnosis, (ii) the TBI occurred before the mental health diagnosis, or (iii) the TBI and mental health diagnosis occurred within the same calendar year. Self-reported TBIs were included if participants reported a lifetime TBI at baseline recruitment assessment and either had no psychiatric diagnosis or their diagnosis occurred in or after 2010 (the latest possible year of baseline assessment). These temporal restrictions ensured that TBI did not occur after the onset of mental health outcomes. Importantly, no assumptions were made regarding whether TBI preceded the onset of neurological disease; analyses only required that both TBI and neurological conditions occurred before any psychiatric outcomes.

To examine the combined influence of TBI and any neurological disease on subsequent psychiatric outcomes, all participants were classified into four mutually exclusive exposure groups: (1) no TBI and no neurological condition, (2) TBI only, (3) neurological condition only, and (4) both TBI and a neurological condition. Mixed-effects logistic regression models were used to compare the odds of mood disorders, anxiety disorders, psychotic disorders, and any mental health disorder across these groups, using the group with neither TBI nor neurological disease as the reference category. Models included study site as a random effect to account for a nested structure, and results were reported as unadjusted and adjusted odds ratios (ORs; 95% CIs). Adjusted models controlled for sex, age, race/ethnicity, socioeconomic status, and alcohol intake at baseline.

To assess condition-specific risk, three analytic cohorts were constructed, consisting of individuals with dementia, stroke, or Parkinson’s disease. Within each cohort, TBI was the exposure and mental health conditions were the outcomes of interest. Only cases in which the neurological condition occurred before or in the same year as the onset of any psychiatric condition were included, and no maximum follow-up period was imposed. Mixed-effects logistic regression models (with site as a random effect) were used to estimate unadjusted and adjusted ORs for each psychiatric outcome within each neurological population.

Kaplan-Meier probability estimates (with 95% CIs) and survival curves were used to summarise and illustrate the cumulative incidence of psychiatric problems with and without TBI up to 10 years following diagnosis of dementia, stroke or Parkinson’s disease. Differences in survival distributions between participants with and without TBI were assessed using the log-rank test. Cox proportional hazards regression was used to estimate the association between TBI and risk of mental health problems in each neurological condition, with unadjusted and adjusted hazard ratios (HR) and corresponding 95% CIs reported. The proportional hazards assumption was checked visually and by review of the Schoenfeld global test. All adjusted survival models incorporated an individual frailty term to account for random effects at the participant level. All analysis was conducted using *R* version 4.5.1. (R Core Team, 2020). The full code for the analysis is available in full in an online archive: https://github.com/GraceRevill/biobank-neuropsychiatric-outcomes.

## Results

### Sample description

In the full UK Biobank cohort (N = 502,154), 12,169 participants had at least one ICD-10-coded TBI, and 1,421 reported a lifetime TBI at baseline assessment, with 215 individuals meeting both criteria. After applying temporal inclusion rules, requiring that TBI and neurological diagnoses occurred before the onset of any mental health disorder, the final analytic sample comprised 25,504 participants with a neurological disorder, including 2,017 individuals with TBI (Figure 1). Of these, 1,891 were identified through ICD-coded records, 101 through self-report, and 25 through both sources.

**Figure 1.**
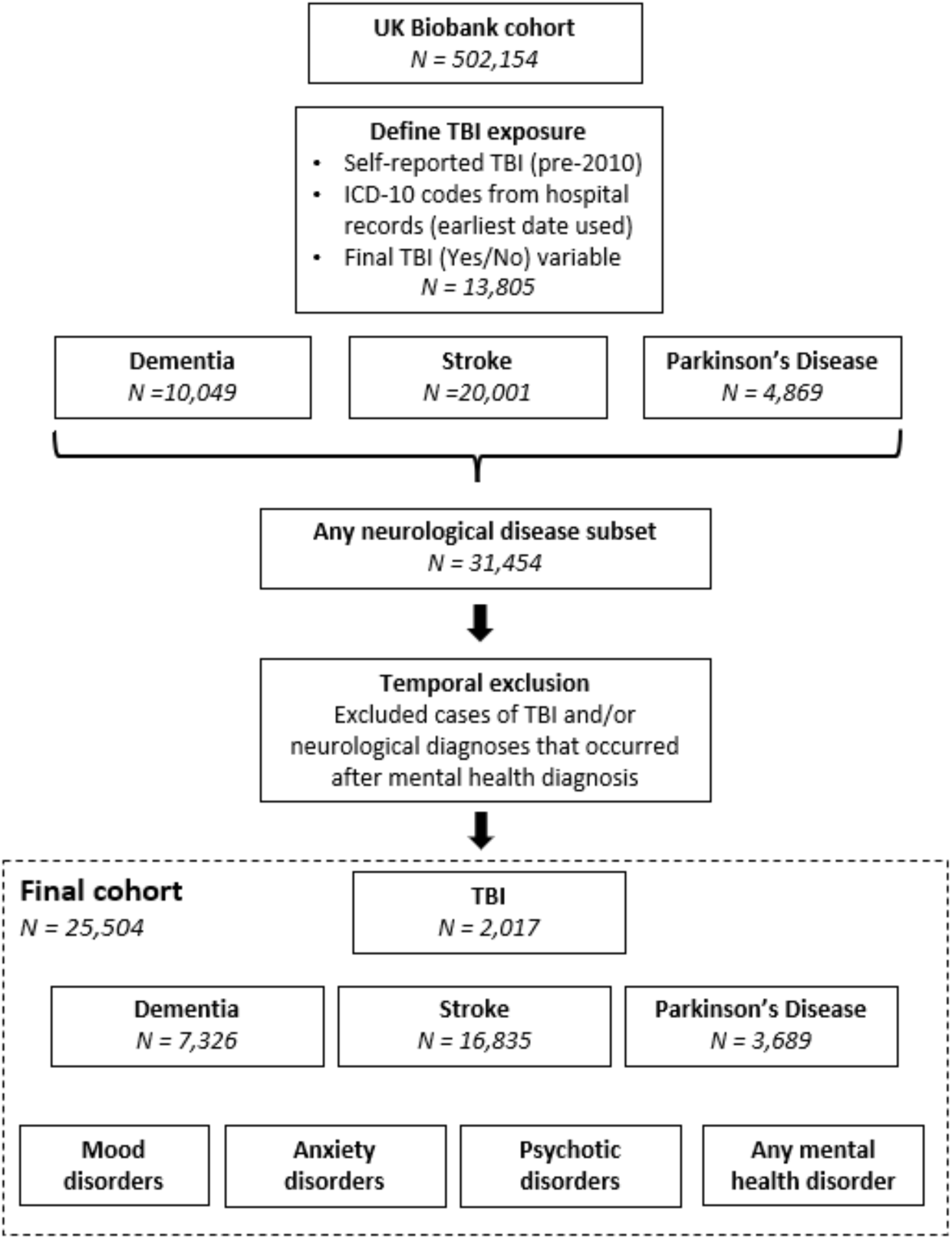
Flow of participants through the cohort selection process.

Baseline demographic and clinical characteristics for the final analytic cohort are presented in Table 1. Age, sex distribution, ethnicity, socioeconomic status, and alcohol intake patterns were broadly comparable across exposure groups. Levels of missing data were low, with most variables complete or containing fewer than 5% missing values. Descriptive statistics for the full Biobank cohort are provided in Supplementary Table 2, and the distribution of neurological conditions by TBI status is reported in Supplementary Table 3.

**Table 1.** Demographics and descriptive statistics for the final sample. ^1^n (%); Median (IQR)

|  | *Overall*<br>*N = 25,504* | *TBI*<br>*N = 2,017* | *Dementia*<br>*N = 7,326* | *Stroke*<br>*N= 16,835* | *Parkinson's*<br>*N= 3,689* |
| --- | --- | --- | --- | --- | --- |
| *Sex* |  |  |  |  |  |
| Female | 10,780 (42%) | 745 (37%) | 3,309 (45%) | 6,913 (41%) | 1,332 (36%) |
| Male | 14,724 (58%) | 1,272 (63%) | 4,017 (55%) | 9,922 (59%) | 2,357 (64%) |
| *Age at baseline<sup>1</sup>* | 64 (59, 67) | 65 (60, 67) | 66 (62, 68) | 63 (57, 66) | 64 (60, 67) |
| *Race and ethnicity* |  |  |  |  |  |
| Asian/Asian British | 507 (2.0%) | 44 (2.2%) | 116 (1.6%) | 368 (2.2%) | 64 (1.7%) |
| Black(Caribbean/African) | 402 (1.6%) | 18 (0.9%) | 115 (1.6%) | 288 (1.7%) | 42 (1.1%) |
| Mixed/multiple | 98 (0.4%) | 9 (0.4%) | 24 (0.3%) | 66 (0.4%) | 14 (0.4%) |
| Other | 167 (0.7%) | 11 (0.5%) | 39 (0.5%) | 112 (0.7%) | 23 (0.6%) |
| White | 24,167 (95%) | 1,919 (96%) | 6,974 (96%) | 15,900 (95%) | 3,523 (96%) |
| Unknown | 163 | 16 | 58 | 101 | 23 |
| *Townsend Deprivation* | -1.9 (-3.5, 1.1) | -1.5 (-3.3, 1.7) | -1.9 (-3.6, 1.0) | -1.7 (-3.4, 1.4) | -2.4 (-3.8, 0.2) |
| Unknown | 69 | 5 | 23 | 43 | 11 |
| *Alcohol intake frequency* |  |  |  |  |  |
| Daily/almost daily | 5,472 (22%) | 480 (24%) | 1,517 (21%) | 3,616 (22%) | 832 (23%) |
| 3/4 times a week | 5,079 (20%) | 387 (19%) | 1,442 (20%) | 3,332 (20%) | 786 (21%) |
| 1/2 a week | 6,067 (24%) | 443 (22%) | 1,684 (23%) | 4,031 (24%) | 877 (24%) |
| 1-3 times a month | 2,512 (9.9%) | 177 (8.8%) | 745 (10%) | 1,662 (9.9%) | 345 (9.4%) |
| Special occasions | 3,317 (13%) | 253 (13%) | 978 (13%) | 2,206 (13%) | 424 (12%) |
| Never | 2,949 (12%) | 262 (13%) | 918 (13%) | 1,925 (11%) | 408 (11%) |
| Unknown | 108 | 15 | 42 | 63 | 17 |

The proportion and percentage of psychiatric disorders among participants with dementia, stroke, or Parkinson’s disease, stratified by TBI history, are summarised in Table 2. Across all neurological conditions, those with TBI exhibited higher prevalence of mood, anxiety, psychotic, and any mental health disorder compared with those without TBI. The overall prevalence of psychiatric disorders in the full Biobank sample, stratified by TBI history and neurological status, is shown in Supplementary Table 4.

**Table 2.** Proportion and percentage of mood, anxiety, and psychotic disorders in individuals with and without TBI, stratified by neurological condition. TBI = traumatic brain injury.

|  | **TBI** |  |
| --- | --- | --- |
|  | *Yes*<br>*N = 2,017* | *No*<br>*N = 23,487* |
| Mood disorders | 262 (13%) | 1,907 (8.1%) |
| Anxiety disorders | 148 (7.3%) | 1,420 (6.0%) |
| Psychotic disorders | 19 (0.9%) | 84 (0.4%) |
| Any mental health disorder | 343 (17%) | 2,842 (12%) |
|  | **Dementia** |  |
|  | *TBI*<br>*N = 868* | *No TBI*<br>*N = 6,458* |
| Mood disorders | 86 (9.9%) | 514 (8%) |
| Anxiety disorders | 57 (6.6%) | 354 (5.5%) |
| Psychotic disorders | 5 (0.6%) | 40 (0.6%) |
| Any mental health disorder | 123 (14.2%) | 766 (11.9%) |
|  | **Stroke** |  |
|  | *TBI*<br>*N = 1,161* | *No TBI*<br>*N = 15,674* |
| Mood disorders | 164 (14.1%) | 1,253 (8%) |
| Anxiety disorders | 79 (6.8%) | 914 (5.8%) |
| Psychotic disorders | 11 (0.9%) | 37 (0.2%) |
| Any mental health disorder | 202 (17.4%) | 1,829 (11.7%) |
|  | **Parkinson's Disease** |  |
|  | *TBI*<br>*N = 389* | *No TBI*<br>*N = 3,300* |
| Mood disorders | 52 (13.4%) | 287 (8.7%) |
| Anxiety disorders | 33 (8.5%) | 238 (7.2%) |
| Psychotic disorders | 5 (1.3%) | 23 (0.7%) |
| Any mental health disorder | 69 (17.7%) | 457 (13.8%) |

### Association between TBI, neurological disease, and subsequent mental health outcomes

When TBI and neurological diagnoses were combined into a four-level exposure variable, all three groups with an exposure (TBI only, neurological disease only, and TBI + neurological disease) demonstrated significantly higher odds of mental health disorders compared with individuals with neither condition, even after adjustment for covariates (Table 3). The greatest risk was observed among participants with both TBI and a neurological disorder, who showed markedly elevated odds of mood, anxiety, and overall mental health disorders relative to those with either condition alone.

**Table 3.** Association between traumatic brain injury, neurological condition, and mental health outcomes. TBI = traumatic brain injury. Neuro = neurological condition (dementia, stroke or Parkinson’s disease).

|  | *Unadjusted*<br>*OR (95% CI)* | *Adjusted*<br>*OR (95% CI)* |
| --- | --- | --- |
| *Mood disorders* |  |  |
| Neuro only | 1.89 (1.83 – 1.95) | 2.07 (2.01 – 2.14) |
| TBI only | 2.04 (1.95 – 2.14) | 2.08 (1.99 – 2.19) |
| Neuro + TBI | 3.56 (3.29 – 3.85) | 3.97 (3.67 – 4.31) |
| *Anxiety disorders* |  |  |
| Neuro only | 1.76 (1.70 – 1.82) | 1.84 (1.77 – 1.90) |
| TBI only | 1.75 (1.66 – 1.85) | 1.82 (1.72 – 1.92) |
| Neuro + TBI | 2.69 (2.45 – 2.95) | 2.85 (2.60– 3.14) |
| *Any mental health disorder* |  |  |
| Neuro only | 1.88 (1.83 – 1.93) | 2.03 (1.97 – 2.08) |
| TBI only | 1.93 (1.85 – 2.01) | 1.99 (1.90 – 2.08) |
| Neuro + TBI | 3.37 (3.13 – 3.63) | 3.70 (3.43 – 4.00) |

Although psychotic disorders were not included in Table 2 due to their low prevalence, the pattern of associations closely mirrored that of the other mental health outcomes. Compared with participants with neither TBI nor neurological disease, the adjusted odds of psychotic disorder were significantly higher in those with TBI only (OR = 3.17, 95% CI 2.67–3.75) and in those with neurological disease only (OR = 3.45, 95% CI 3.07–3.88). The largest effect was observed in participants with both TBI and a neurological disorder (OR = 7.93, 95% CI 6.39–9.85).

### Association between TBI and mental health outcomes in specific neurological condition populations

#### Dementia: association between TBI and psychiatric complications

As shown in Table 4, individuals with a history of TBI had higher odds of developing mood and anxiety disorders at any point after dementia diagnosis compared with those without TBI. TBI was also associated with elevated, though not statistically significant, odds of psychotic disorders (unadjusted OR = 1.69, 95% CI 0.87–3.30; adjusted OR = 1.61, 95% CI 0.82–3.15). Overall, lifetime TBI was associated with greater odds of any mental health disorder following dementia, and these associations persisted after adjustment for demographic and lifestyle covariates.

**Table 4.** Association between psychiatric disorders and lifetime TBI in people with neurological conditions. TBI = traumatic brain injury. OR = odds ratio.

|  | *Unadjusted*<br>*OR (95% CI)* | *Adjusted*<br>*OR (95% CI)* |
| --- | --- | --- |
| *Dementia* |  |  |
| Mood disorders | 2.15 (1.75 – 2.64) | 2.10 (1.70 – 2.59) |
| Anxiety disorders | 1.91 (1.53 – 2.39) | 1.97 (1.57 – 2.48) |
| Any mental health disorder | 1.99 (1.65 – 2.40) | 1.96 (1.62 – 2.37) |
| *Stroke* |  |  |
| Mood disorders | 1.72 (1.48 – 2.01) | 1.76 (1.50 – 2.06) |
| Anxiety disorders | 1.02 (0.83 – 1.25) | 1.09 (0.89 – 1.34) |
| Any mental health disorder | 1.55 (1.34 – 1.78) | 1.59 (1.37 – 1.83) |
| *Parkinson's disease* |  |  |
| Mood disorders | 2.65 (2.10 – 3.36) | 2.70 (2.13 – 3.43) |
| Anxiety disorders | 1.84 (1.40 – 2.42) | 1.96 (1.48 – 2.58) |
| Any mental health disorder | 2.2 (1.77 – 2.74) | 2.27 (1.82 – 2.83) |

Over a follow-up period of up to 10 years, Kaplan-Meier curves demonstrated higher cumulative incidence of mood disorders among participants with dementia who had a history of TBI compared with those without (log-rank test, *p* = 0.002; Figure 2). Consistent with this, Cox proportional hazards models indicated that TBI was associated with an increased risk of mood disorders following dementia (Table 5). For anxiety disorders, the difference between groups was not statistically significant in the Kaplan-Meier analysis (log-rank test, *p* = 0.075; Figure 2). However, the adjusted Cox model suggested a modest but significant association between TBI and anxiety disorders after dementia (Table 5). For any mental health disorder, cumulative incidence was higher among dementia participants with TBI (log-rank test, *p* = 0.0086; Figure 2), and TBI remained significantly associated with increased risk in Cox models (Table 5). The proportional hazards assumption was evaluated using Schoenfeld residuals and was not violated for any adjusted dementia model (Supplementary Table 5).

**Figure 2.**
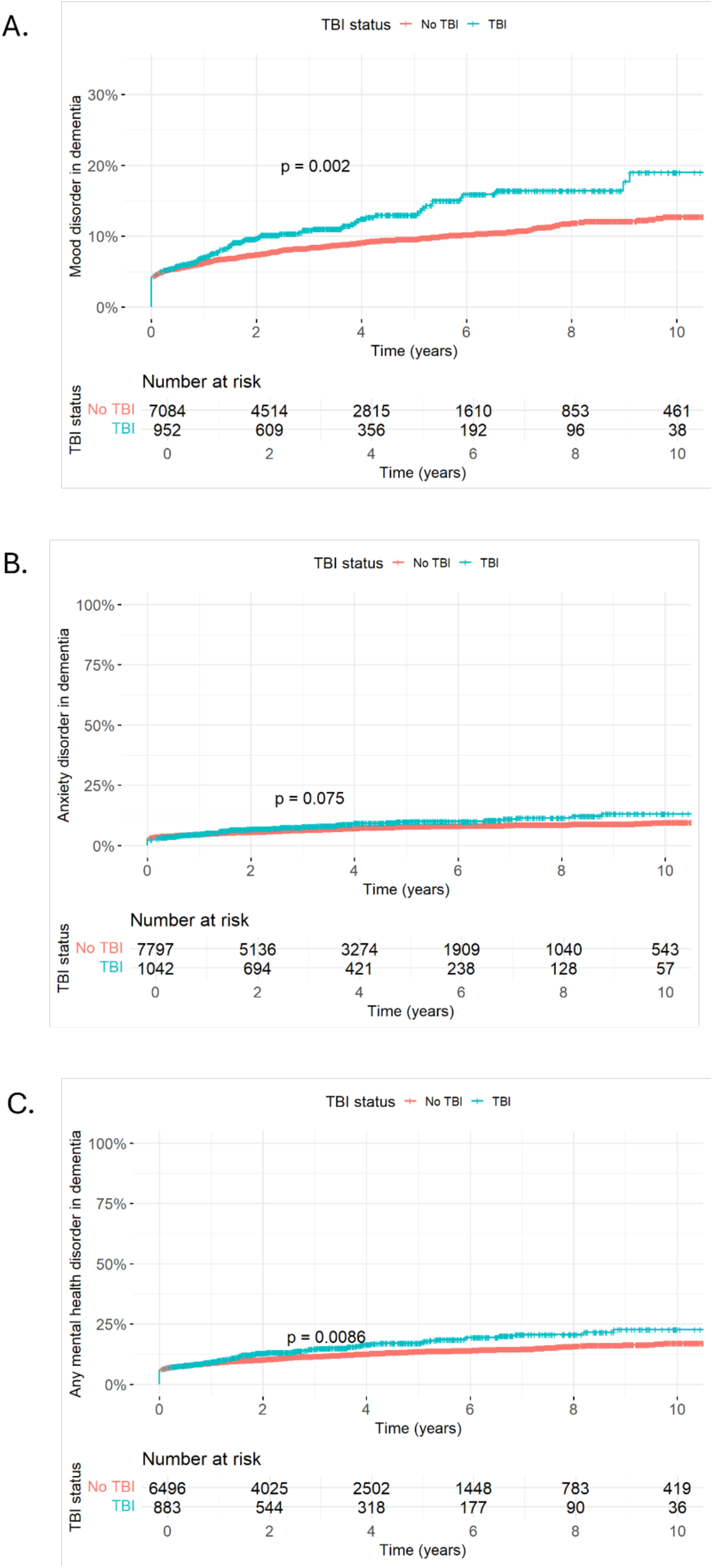
The cumulative incidence of A) mood B) anxiety disorders and C) any mental health disorder in participants with dementia reporting and not reporting TBI

**Table 5.** Hazard ratio estimates for psychiatric disorder risk after TBI in dementia, stroke and Parkinson’s disease. HR = Hazard ratio.

|  | *Unadjusted*<br>*HR (95% CI)* | *Adjusted*<br>*HR (95% CI)* |
| --- | --- | --- |
| *Dementia* |  |  |
| Mood disorders | 1.36 (1.12–1.67) | 1.40 (1.14–1.71) |
| Anxiety disorders | 1.22 (0.98–1.53) | 1.31 (1.04–1.64) |
| Any mental health disorder | 1.27 (1.06–1.52) | 1.31 (1.09–1.58) |
| *Stroke* |  |  |
| Mood disorders | 1.89 (1.64–2.19) | 1.86 (1.60–2.15) |
| Anxiety disorders | 1.16 (0.95–1.41) | 1.20 (0.98–1.46) |
| Any mental health disorder | 1.67 (1.47–1.91) | 1.66 (1.45–1.90) |
| *Parkinson's disease* |  |  |
| Mood disorders | 1.73 (1.38–2.18) | 1.77 (1.40–2.23) |
| Anxiety disorders | 1.23 (0.94–1.62) | 1.30 (0.99–1.71) |
| Any mental health disorder | 1.43 (1.16–1.77) | 1.47 (1.18–1.81) |

#### Stroke: association between TBI and psychiatric complications

As shown in Table 4, individuals with a history of TBI had higher odds of developing mood disorders at any point after stroke compared with those without TBI. TBI was also associated with increased odds of psychotic disorders (unadjusted OR = 1.98, 95% CI 1.07–3.66; adjusted OR = 2.05, 95% CI 1.10–3.81). For anxiety disorders, the association was not statistically significant. Overall, lifetime TBI was linked to greater odds of any mental health disorder following stroke, and these associations remained significant after adjustment for covariates.

Over a follow-up period of up to 10 years, the Kaplan-Meier curves demonstrated higher cumulative incidence of mood disorders among stroke participants with TBI compared to those without (log-rank test, *p* < 0.0001; Figure 3). Consistent with this, Cox proportional hazards models indicated that TBI was associated with an increased risk of mood disorders following stroke (Table 5). For anxiety disorders, the Kaplan-Meier curves did not show a statistically significant difference between TBI and non-TBI groups (log-rank test, *p* = 0.15; Figure 3). However, the adjusted Cox model suggested a small but non-significant elevation in risk among those with TBI (Table 5). For any mental health disorder, cumulative incidence was substantially higher in the TBI group (log-rank test, *p* < 0.0001; Figure 3), and the adjusted Cox model confirmed that TBI was associated with an increased hazard of any mental health disorder following stroke (Table 5).The proportional hazards assumption was tested using Schoenfeld residuals and was not violated for any stroke model (Supplementary Table 5).

**Figure 3.**
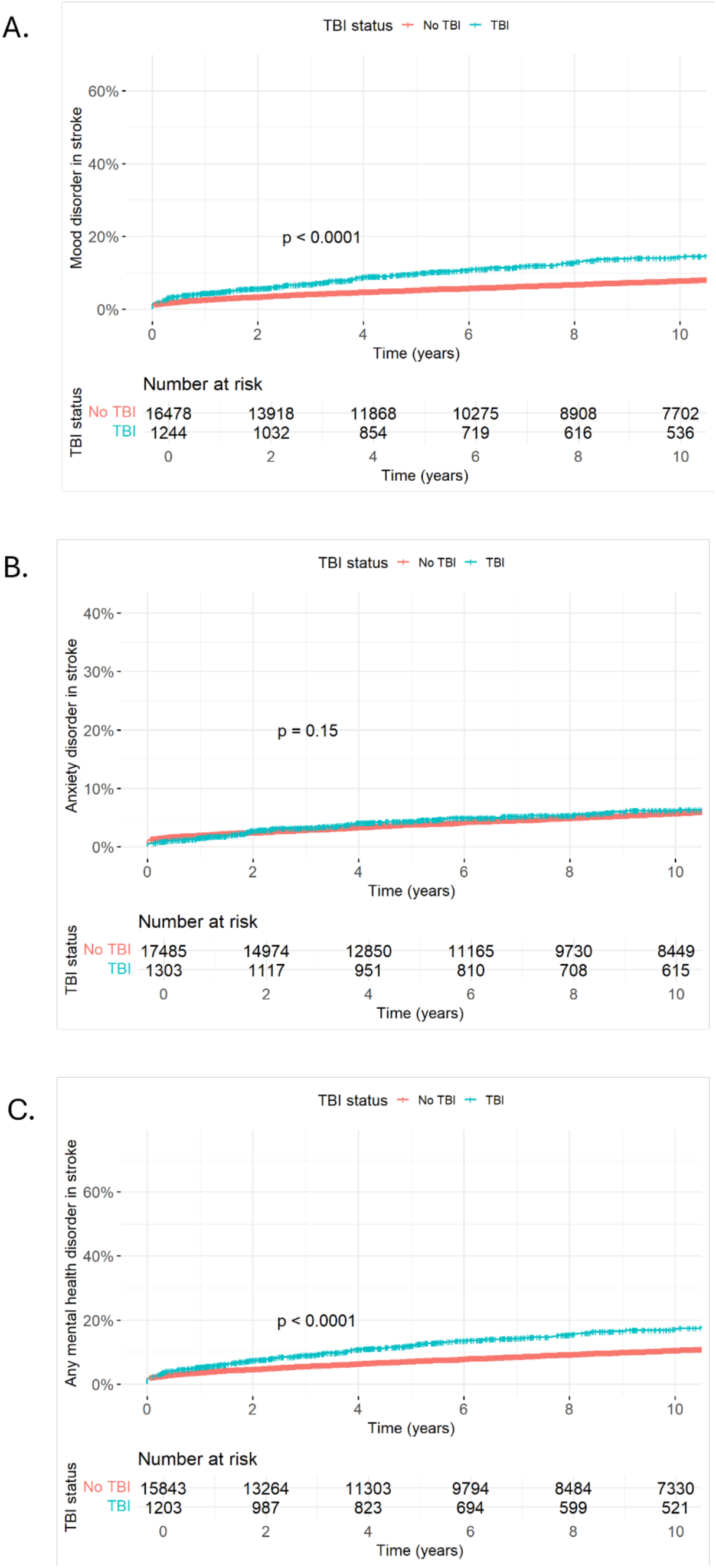
The cumulative incidence of A) mood B) anxiety disorders and C) any mental health disorder in participants with stroke reporting and not reporting TBI

#### Parkinson’s disease: association between TBI and psychiatric complications

As shown in Table 4, individuals with a history of TBI had higher odds of developing mood and anxiety disorders at any point after Parkinson’s disease compared with those without TBI. The number of psychotic disorder cases was too small to estimate reliable odds ratios (unadjusted OR = 3.06, 95% CI 1.70–5.52; adjusted OR not estimable). Overall, lifetime TBI was linked to greater odds of any mental health disorder following Parkinson’s disease, and these associations remained significant after covariate adjustment.

Over a follow-up of up to 10 years, Kaplan-Meier estimates demonstrated higher cumulative incidence of mood disorders among Parkinson’s disease participants with TBI compared to those without (log-rank test, *p* < 0.0001; Figure 4). The corresponding Cox model indicated that TBI was associated with substantially increased hazard of mood disorders (Table 5). For anxiety disorders, cumulative incidence did not differ significantly by TBI status (log-rank test, *p* = 0.12; Figure 4), however, the adjusted Cox model suggested a small but non-significant elevation in risk among those with TBI (Table 5). For any mental health disorder, TBI was associated with markedly higher cumulative incidence (log-rank test, *p* = 0.00075; Figure 4) and a significantly increased hazard of developing any mental health disorder after Parkinson’s disease (Table 5). The proportional hazards assumption was evaluated using Schoenfeld residuals and was not violated for any adjusted Parkinson’s disease model (Supplementary Table 5).

**Figure 4.**
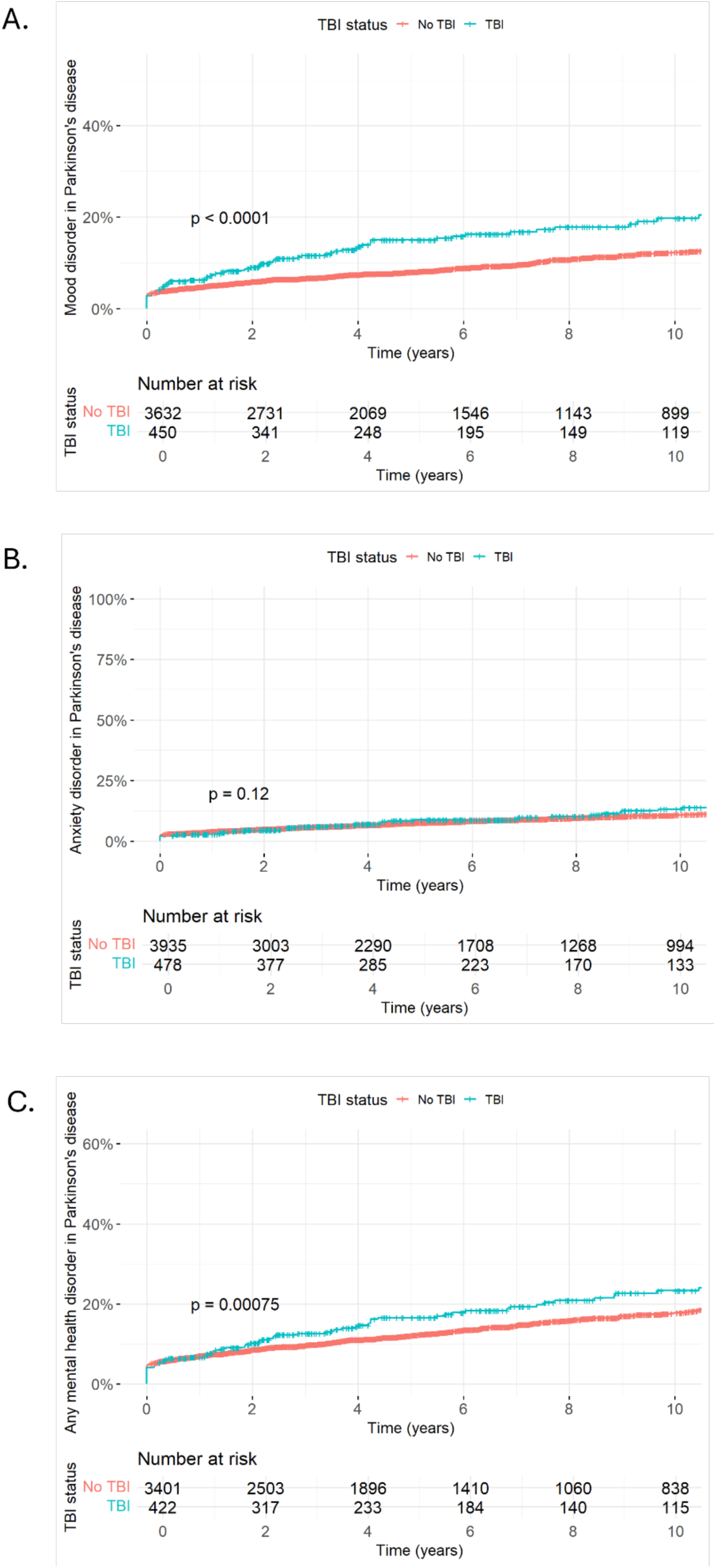
The cumulative incidence of A) mood B) anxiety disorders and C) any mental health disorder in participants with Parkinson’s disease reporting and not reporting TBI

## Discussion

Using data from the UK Biobank, a large prospective cohort of over 500,000 adults, this study provides evidence that TBI is associated with increased psychiatric vulnerability following the onset of major neurological conditions. Across dementia, stroke, and Parkinson’s disease, individuals with TBI consistently showed elevated odds of developing mental health problems, with mood disorders emerging as the most robust outcome.

When TBI and neurological conditions were examined jointly using a four-level exposure model, all three exposed groups (TBI only, neurological disease only, and TBI + neurological disease) demonstrated significantly greater odds of psychiatric disorders compared with those with neither condition. The combined exposure group showed the largest effects, including markedly increased odds of mood, anxiety, and overall mental health disorders. Psychotic disorders, although rare, followed a similar pattern, with the highest odds observed in those with both TBI and neurological disease, although the few cases meant the estimate was not statistically reliable. Mood disorders showed the strongest associations. This graded pattern of risk is consistent with an additive, and potentially synergistic, relationship in which TBI increases psychiatric vulnerability beyond that associated with neurological disease alone, rather than acting solely as a precursor to neurological illness.

The disease-specific analyses further revealed that the nature and consistency of associations vary across neurological conditions. In dementia, TBI was associated with higher odds of mood and anxiety disorders and with elevated, but not statistically significant, odds of psychotic disorders. Survival analyses indicated that TBI confers increased 10-year risk of mood disorders and any mental health disorder following dementia diagnosis, whereas evidence for anxiety was weaker. In stroke, TBI was associated with higher odds of mood and psychotic disorders, but not anxiety, and with increased 10-year cumulative incidence of mood and any mental health disorder. In Parkinson’s disease, TBI was associated with elevated odds of mood and anxiety disorders, although limited case numbers prevented adjusted psychotic disorder estimates. Among the neurological conditions examined, Parkinson’s disease showed the strongest overall association between TBI and subsequent mental health disorders. Over 10 years of follow-up, mood and overall mental health disorders also showed greater cumulative incidence in individuals with TBI, while anxiety disorder risk was not significantly increased. These condition-specific patterns suggest that psychiatric risk following TBI is not uniform across neurological diseases but instead may depend on interactions between prior injury-related vulnerability and disease-specific neuropathology.

Conceptually, these findings are most consistent with a “force multiplier” model, in which TBI, regardless of its timing relative to neurological disease, amplifies psychiatric vulnerability in the context of neurological pathology. Such interactive effects are consistent with research on multimorbidity and comorbidity, where co-occurring conditions can interact to produce outcomes that exceed the sum of individual disease effects (Harrison et al., 2021). The persistence of elevated psychiatric risk over a decade further supports the presence of a durable alteration in vulnerability rather than a transient effect related to diagnosis or healthcare contact. This interpretation contrasts with a simpler mediation model in which elevated psychiatric risk would be fully explained by neurological disease itself, with no additional contribution from TBI. This explanation is unlikely to fully account for the findings, as the presence of TBI was associated with additional psychiatric risk beyond that observed for neurological disease alone, with patterns that varied by neurological condition and psychiatric outcome.

Although the risk-amplifier model provides the most coherent account of the findings, it is important to evaluate whether simpler explanations could account for the observed pattern. One possibility is that TBI represents a non-specific marker of poorer overall health or greater disease severity, such that individuals with TBI simply experience worse outcomes across domains when neurological disease develops (Hanafy et al., 2021; Xiong et al., 2019). Although plausible, this explanation is not fully supported by the findings. Notably, the absence of increased anxiety risk following stroke, despite associations with mood disorders and with anxiety in other neurological conditions, argues against a non-specific severity effect and instead supports the idea that the characteristic neuropathological features of TBI interact with disease-specific brain changes to selectively increase psychiatric risk. The neuropathological signature of TBI, characterised by disruption to prefrontal-limbic connectivity and frontostriatal circuits involved in emotional appraisal, regulation, and reward, overlaps selectively with the circuits most affected in dementia and Parkinson’s disease, but less consistently with the lesion distributions typical of stroke, which vary substantially (Bryant et al., 2022; Fischer et al., 2021; Newsome et al., 2013; van der Horn et al., 2016). In addition, diffuse axonal injury may produce widespread but subtle network disruption that disproportionately affects affective and regulatory processes, increasing vulnerability to psychiatric disorder (Bryant et al., 2022; Hayes et al., 2016; Mesfin et al., 2025). Consistent with this interpretation, prior work has demonstrated that different neurological disorders show distinct mental health profiles, supporting the expectation that psychiatric outcomes vary according to underlying pathophysiology rather than reflecting a uniform effect of disease severity (Burchill et al., 2025). Although this work did not examine TBI specifically, it provides a useful framework for anticipating differential psychiatric outcomes based on neuropathological mechanisms. Beyond these biological mechanisms, psychosocial and functional consequences of both TBI and neurological disease are also likely to contribute to elevated psychiatric risk. Reduced independence, social isolation, loss of meaningful roles, and cumulative health burden are common following both conditions and may exacerbate affective vulnerability over time (Backhouse et al., 2018; Bombardier et al., 2010; Cnossen et al., 2017; Maas et al., 2022; Perepezko et al., 2019). Importantly, individuals with TBI and neurological disease may have pre-existing functional limitations or reduced coping capacity, such that the psychosocial impact of neurological diagnosis is experienced more acutely and persists over time.

Together, these mechanisms may be conceptualised as reflecting a reduction in what could be termed ‘neuropsychiatric reserve’, by analogy with the concept of cognitive reserve in dementia research (Stern, 2012). In this context, neuropsychiatric reserve refers to the capacity to maintain affective stability and emotional regulation in the face of brain pathology or psychosocial stress. Although this construct is not yet formally defined, it provides a useful framework for understanding why individuals with TBI may be less able to compensate for the combined neurobiological and psychosocial challenges posed by neurological disease. Future research is needed to determine whether such a reserve can be empirically measured and how it relates to established concepts of resilience and cognitive reserve.

Across all neurological conditions, mood disorders emerged early and persisted throughout the 10-year follow-up, indicating a stable and enduring vulnerability that was strongest when TBI and neurological disease co-occurred. This makes mood disorders the clearest clinical manifestation of the proposed ‘risk-amplifier’ effect. Rather than reflecting transient distress around diagnosis, the findings suggest long-term alterations in affective regulation associated with TBI that reduce capacity to adapt to neurological pathology or accumulating psychosocial stressors. Neural systems central to mood regulation, including prefrontal, limbic, and frontostriatal networks, are particularly vulnerable to both traumatic injury and the neuropathological processes characteristic of dementia, stroke, and Parkinson’s disease, providing a plausible biological substrate for this persistent risk (Mavroudis et al., 2022; Park et al., 2019). This aligns with extensive research demonstrating that depression is among the most common and persistent sequelae of TBI, even at mild levels of severity (Howlett et al., 2022; Ogonah et al., 2025), and one of the most prevalent psychiatric conditions across major neurological disorders in later life (Aarsland et al., 2009; Cloak et al., 2025; Hackett et al., 2014; Valiengo et al., 2016). Psychosocial factors such as functional limitation, social isolation, and cumulative health burden are also likely to compound this vulnerability over time, further reinforcing mood disorders as the most consistent psychiatric outcome in this population (Backhouse et al., 2018; Bombardier et al., 2010; Cnossen et al., 2017; Maas et al., 2022; Perepezko et al., 2019).

In contrast to mood disorders, anxiety showed greater heterogeneity across neurological conditions and over time. Although lifetime TBI was significantly associated with increased odds of anxiety disorders in dementia and Parkinson’s disease when considering any time after diagnosis, this pattern was not reflected in the 10-year cumulative incidence curves. Kaplan-Meier analysis indicates minimal early separation between TBI and non-TBI groups, with the curves only beginning to diverge toward the later years of follow-up. This pattern suggests that anxiety develops or is detected later in the disease course, particularly as functional decline increases and neuropsychiatric symptoms become more clinically prominent. For example, mental health symptoms such as anxiety are frequently under-recognised in dementia due to diagnostic overshadowing, whereby cognitive decline or behavioural disturbance dominate clinical attention, reducing the likelihood that anxiety is identified or recorded in the early stages of disease (Dunn et al., 2022). No association between TBI and anxiety was observed following stroke, either in odds or long-term incidence. This divergence may reflect fundamental differences in disease trajectories: whereas dementia and Parkinson’s disease involve progressive decline that may sustain or amplify anxiety, stroke is typically non-degenerative, with anxiety often peaking early (Campbell Burton et al., 2013; Emmady et al., 2025; Mack & Marsh, 2017). In addition, the heterogeneity of stroke lesions may dilute any added effect of TBI, as strokes can involve regions strongly or only weakly linked to anxiety, so symptom emergence depends heavily on lesion location and size. This variability may overshadow contributions from TBI. In contrast, the more predictable involvement of frontostriatal and limbic circuits in dementia and Parkinson’s disease, networks also often disrupted by TBI, may increase the likelihood that TBI-related vulnerability to anxiety becomes clinically evident in these groups (Brown et al., 2023; Cotta Ramusino et al., 2024)..

This study has several important strengths. The use of a large, well-characterised cohort with detailed data on neurological diagnoses, psychiatric outcomes, and TBI exposure allowed us to address questions that smaller studies cannot. By applying strict temporal inclusion rules, ensuring that TBI and neurological conditions preceded mental health outcomes, we were able to rigorously evaluate whether lifetime TBI contributes to subsequent psychiatric risk in neurological populations. The prospective design and long follow-up enabled us to examine cumulative psychiatric risk up to 10 years after neurological diagnosis. Furthermore, by analysing both overall odds and long-term incidence of mood, anxiety, and psychotic disorders, this study addresses a critical gap in the literature, which has typically examined TBI, neurological disease, and psychiatric outcomes in isolation. Considering three major neurological conditions simultaneously enabled us to identify both condition-specific patterns of vulnerability and consistent findings, particularly the elevated risk of mood disorders, that cut across diagnoses. Importantly, although the association between TBI and later psychiatric morbidity is well established, older adults have been strikingly underrepresented in previous research. Much of the existing evidence is derived from younger or middle-aged samples, leaving substantial uncertainty about whether these findings generalise to ageing populations. This is a critical gap, given that older adults now represent the group with the highest rates of TBI-related hospitalisation and mortality (Maas et al., 2022; Thompson et al., 2006). The present study therefore provides timely and clinically relevant evidence by examining lifetime TBI within a large cohort of older adults. By capturing psychiatric outcomes after neurological onset, this work offers new insight into long-term vulnerability in later life and highlights the importance of recognising TBI history as a potentially enduring risk factor for mental health problems in ageing neurological populations.

However, it is important to note some limitations of this study. First, because the number of TBI cases meeting strict temporal criteria was limited, we were unable to stratify by injury characteristics or examine more refined temporal relationships. As a result, TBI cases were grouped solely on the basis that the injury occurred before the onset of mental health problems, regardless of whether it occurred before or after the neurological diagnosis. To maximise case inclusion, TBI events occurring in the same calendar year as a mental health diagnosis were retained, introducing a small possibility that some psychiatric outcomes may have preceded either the TBI or the neurological condition. Second, the psychiatric outcomes reflect recorded diagnoses, which likely underestimate the true burden of mental health symptoms. Many individuals experience significant distress without receiving a formal diagnosis, and psychiatric conditions are often under-recognised in people with neurological disorders (Aarsland et al., 2009; Hackett et al., 2014; Lyketsos et al., 2000). In addition, diagnostic overshadowing, where new or emerging psychiatric symptoms are misattributed to the underlying neurological condition, further contributes to missed or delayed identification of mental health problems in this population (Dunn et al., 2022). As such, the present estimates likely understate the true burden of psychiatric symptoms in these populations. In addition, although analyses were restricted to those whose mental health diagnosis occurred after TBI and/or neurological disease, some participants may have had earlier symptoms that were only identified once they engaged with healthcare services due to their neurological condition. Third, several aspects of TBI exposure could not be characterised: severity, mechanism of injury, and whether participants experienced multiple TBIs were unavailable, preventing examination of dose-response or cumulative injury effects. The relatively low number of ICD-10-defined TBI cases in this cohort suggests that some injuries may have been unreported and/or been represented in primary care diagnoses which were not analysed here. Self-reported TBI is inherently heterogeneous and may be subject to recall bias (Perry et al., 2016), and the small number of cases meant that ICD-coded and self-reported TBIs could not be analysed separately, limiting the ability to compare these exposure sources.

Fourth, although covariates were carefully selected and adjusted for, residual confounding cannot be ruled out. Individuals with TBI may differ systematically from those without TBI on unmeasured factors such as premorbid mental health, lifetime socioeconomic adversity, substance use, chronic pain, or personality traits, all of which could contribute to later psychiatric risk. However, the persistence of associations over long follow-up, their differential pattern across neurological conditions and psychiatric outcomes, and their survival beyond adjustment for measured sociodemographic and health-related factors suggest that unmeasured confounding alone is unlikely to fully account for the observed findings. Fifth, the small number of psychotic disorder cases, particularly in Parkinson’s disease, prevented survival analyses for this outcome, limiting our ability to fully characterise associations. Finally, the UK Biobank cohort is healthier and less socioeconomically deprived than the general population, which may affect generalisability (Fry et al., 2017). Individuals with more severe TBI and/or neurological diagnosis may have been less able or likely to participate, creating a “healthy survivor” bias. As a result, the associations observed here may underestimate the true magnitude of psychiatric vulnerability linked to TBI in neurological populations.

Overall, this study shows that lifetime TBI is associated with increased risk of psychiatric disorders in individuals with major neurological disease, with mood disorders demonstrating the most consistent elevation across all conditions, whereas associations with anxiety were weaker and more variable. Taken together, these results suggest that lifetime TBI confers a sustained and clinically meaningful vulnerability to psychiatric morbidity alongside major neurological conditions, although the nature and strength of this risk differ across neurological disorders. These findings underscore the importance of considering TBI history in clinical assessment and care pathways for neurological populations and highlight the need for integrated models of care that address both neurological and psychiatric needs.

## Data Availability

All data produced are available online at: https://github.com/GraceRevill/biobank-neuropsychiatric-outcomes

https://www.ukbiobank.ac.uk/

