## Supplementary material for "Traumatic Brain Injury Increases Risk of Psychiatric Disorders Following Neurological Disease: Evidence from the UK Biobank Cohort"

| **Code** | **Description** |
| --- | --- |
| **S02.01** | Fracture of vault of skull (open) |
| **S02.7** | Multiple fractures involving skull and facial bones |
| **S02.70** | Multiple fractures involving skull and facial bones (closed) |
| **S02.71** | Multiple fractures involving skull and facial bones (open) |
| **S02.8** | Fractures of other skull and facial bones |
| **S02.80** | Fractures of other skull and facial bones (closed) |
| **S02.81** | Fractures of other skull and facial bones (open) |
| **S06.0** | Concussion |
| **S06.00** | Concussion (without open intracranial wound) |
| **S06.01** | Concussion (with open intracranial wound) |
| **S06.1** | Traumatic cerebral oedema |
| **S06.10** | Traumatic cerebral oedema (without open intracranial wound) |
| **S06.11** | Traumatic cerebral oedema (with open intracranial wound) |
| **S06.2** | Diffuse brain injury |
| **S06.20** | Diffuse brain injury (without open intracranial wound) |
| **S06.21** | Diffuse brain injury (with open intracranial wound) |
| **S06.3** | Focal brain injury |
| **S06.30** | Focal brain injury (without open intracranial wound) |
| **S06.31** | Focal brain injury (with open intracranial wound) |
| **S06.4** | Epidural haemorrhage |
| **S06.40** | Epidural haemorrhage (without open intracranial wound) |
| **S06.41** | Epidural haemorrhage (with open intracranial wound) |
| **S06.5** | Traumatic subdural haemorrhage |
| **S06.50** | Traumatic subdural haemorrhage (without open intracranial wound) |
| **S06.51** | Traumatic subdural haemorrhage (with open intracranial wound) |
| **S06.6** | Traumatic subarachnoid haemorrhage |
| **S06.60** | Traumatic subarachnoid haemorrhage (without open intracranial wound) |
| **S06.61** | Traumatic subarachnoid haemorrhage (with open intracranial wound) |
| **S06.7** | Intracranial injury with prolonged coma |
| **S06.70** | Intracranial injury with prolonged coma (without open intracranial wound) |
| **S06.71** | Intracranial injury with prolonged coma (with open intracranial wound) |
| **S06.8** | Other intracranial injuries |
| **S06.80** | Other intracranial injuries (without open intracranial wound) |
| **S06.81** | Other intracranial injuries (with open intracranial wound) |
| **S06.9** | Intracranial injury, unspecified |
| **S06.90** | Intracranial injury, unspecified (without open intracranial wound) |
| **S06.91** | Intracranial injury, unspecified (with open intracranial wound) |
| **S07.9** | Crushing injury of head, part unspecified |
| **S09** | Other and unspecified injuries of head |
| **S09.8** | Other specified injuries of head |
| **S09.9** | Unspecified injury of head |
| **S02.1** | Fracture of base of skull |
| **S02.10** | Fracture of base of skull (closed) |
| **S02.11** | Fracture of base of skull (open) |
| **S02** | Fracture of skull and facial bones |
| **S06** | Intracranial injury |
| **S01.0** | Open wound of scalp |
| **S01.7** | Multiple open wounds of head |
| **S07** | Crushing injury of head |
| **S08** | Traumatic amputation of part of head |
| **S09.7** | Multiple injuries of head |
| **S02.0** | Fracture of vault of skull |
| **S02.00** | Fracture of vault of skull (closed) |

**Supplementary Table 1:** ICD-10 terms used to derive TBI status

|  | ***Overall***  ***N = 502,154*** | ***TBI***  ***N = 13,805*** | | ***Dementia***  ***N = 10,049*** | ***Stroke***  ***N= 20,001*** | ***Parkinson’s***  ***N= 4,869*** |
| --- | --- | --- | --- | --- | --- | --- |
| *Sex* |  | | | | | |
| Female | 273,169 (54%) | 5,672 (41%) | | 4,809 (48%) | 8,590 (43%) | 1,875 (39%) |
| Male | 228,985 (46%) | 8,133 (59%) | | 5,240 (52%) | 11,411 (57%) | 2,994 (61%) |
| *Age at baseline^1^* | 58 (50, 63) | 60 (52, 65) | | 65 (62,68) | 63 (57,66) | 64 (60,67) |
| *Race and ethnicity* |  | | | | | |
| Asian or Asian British | 11,443 (2.3%) | 232 (1.7%) | | 159 (1.6%) | 418 (2.1%) | 85 (1.8%) |
| Black, Caribbean or African | 8,048 (1.6%) | 133 (1.0%) | | 146 (1.5%) | 322 (1.6%) | 56 (1.2%) |
| Mixed/multiple ethnic groups | 2,950 (0.6%) | 77 (0.6%) | | 35 (0.4%) | 86 (0.4%) | 17 (0.4%) |
| Other | 4,553 (0.9%) | 102 (0.7%) | | 59 (0.6%) | 132 (0.7%) | 32 (0.7%) |
| White | 472,385 (95%) | 13,147 (96%) | | 9,573 (96%) | 18,923 (95%) | 4,648 (96%) |
| Unknown | 2,775 | 114 | | 77 | 120 | 31 |
| *Townsend Deprivation Index* | -2.13 (-3.65, 0.55) | -1.44 (-3.32, 1.83) | | -1.86 (-3.50, 1.25) | -1.66 (-3.36, 1.56) | -2.22 (-3.70, 0.58) |
| Unknown | 1,447 | 33 | | 27 | 56 | 18 |
| *Alcohol intake frequency* |  | | | | | |
| Daily/almost daily | 101,716 (20%) | 3,180 (23%) | | 2,022 (20%) | 4,202 (21%) | 1,060 (22%) |
| 3/4 times a week | 115,364 (23%) | 2,844 (21%) | | 1,883 (19%) | 3,825 (19%) | 970 (20%) |
| 1/2 a week | 129,192 (26%) | 3,260 (24%) | | 2,305 (23%) | 4,701 (24%) | 1,137 (23%) |
| 1-3 times a month | 55,813 (11%) | 1,410 (10%) | | 991 (9.9%) | 2,002 (10%) | 454 (9.4%) |
| Special occasions | 57,968 (12%) | 1,581 (12%) | | 1,420 (14%) | 2,749 (14%) | 612 (13%) |
| Never | 40,601 (8.1%) | 1,445 (11%) | | 1,373 (14%) | 2,447 (12%) | 613 (13%) |
| Unknown | 1,500 | 85 | | 55 | 75 | 23 |

**Supplementary Table 2.** Demographics and descriptive statistics for the full UK Biobank sample. ^1^n (%); Median (IQR)

|  | **TBI** | |
| --- | --- | --- |
|  | *Yes*  *N = 13,805* | *No*  *N = 488,349* |
| Dementia | 1,369 (9.9%) | 8,680 (1.8%) |
| Stroke | 1,602 (12%) | 18,399 (3.8%) |
| Parkinson’s Disease | 600 (4.3%) | 4,269 (0.9%) |

**Supplementary Table 3.** Proportion and percentage of neurological conditions among UK Biobank participants with and without a history of traumatic brain injury

|  | | **Total UK Biobank sample** | | |
| --- | --- | --- | --- | --- |
|  | | *N = 502,154* | | |
| Mood disorders | | 65,655 (13%) | | |
| Anxiety disorders | | 45,502 (91%) | | |
| Psychotic disorders | | 2,357 (0.5%) | | |
| Any mental health disorder | | 90,106 (18%) | | |
|  | | **TBI** | | |
|  | | *Yes*  *N = 13,805* | | *No*  *N = 488,349* |
| Mood disorders | | 3,367 (24%) | | 62,288 (13%) |
| Anxiety disorders | | 2,095 (15%) | | 43, 407 (8.9%) |
| Psychotic disorders | | 257 (1.9%) | | 2,100 (0.4%) |
| Any mental health disorder | | 4,234 (31%) | | 85,872 (18%) |
|  | | **Dementia** | | |
|  | | *TBI*  *N = 1,369* | | *No TBI*  *N = 8,680* |
| Mood disorders | | 501 (37%) | | 2,257 (26%) |
| Anxiety disorders | | 304 (22%) | | 1,542 (18%) |
| Psychotic disorders | | 51 (3.7%) | | 195 (2.2%) |
| Any mental health disorder | | 612 (45%) | | 3,000 (35%) |
|  | | **Stroke** | | |
|  | | *TBI*  *N = 1,602* | | *No TBI*  *N = 18,399* |
| Mood disorders | | 502 (31%) | | 3,514 (19%) |
| Anxiety disorders | | 286 (18%) | | 2,274 (12%) |
| Psychotic disorders | | 38 (2.4%) | | 143 (0.8%) |
| Any mental health disorder | | 610 (38%) | | 4,587 (25%) |
|  | **Parkinson’s Disease** | | | |
|  | *TBI*  *N = 524* | | *No TBI*  *N = 4,345* | |
| Mood disorders | 223 (37%) | | 1,055 (25%) | |
| Anxiety disorders | 143 (24%) | | 764 (18%) | |
| Psychotic disorders | 41 (6.8%) | | 129 (3.0%) | |
| Any mental health disorder | 271 (45%) | | 1,435 (34%) | |

**Supplementary Table 4**. Proportion and percentage of mental health disorders in the full UK Biobank cohort stratified by TBI and neurological status

|  | | *P* |
| --- | --- | --- |
| **Dementia** | | |
| *Mood disorders* | |  |
| Unadjusted model | | 0.01 |
| Adjusted Model | | 0.24 |
| *Anxiety disorders* | |  |
| Unadjusted model | | 0.05 |
| Adjusted model | | 0.43 |
| *Any mental health disorder* | |  |
| Unadjusted model | | 0.03 |
| Adjusted model | | 0.29 |
| **Stroke** | | |
| *Mood disorders* | |  |
| Unadjusted model | | 0.48 |
| Adjusted Model | | 0.15 |
| *Anxiety disorders* | |  |
| Unadjusted model | | 0.06 |
| Adjusted model | | 0.94 |
| *Any mental health disorder* | |  |
| Unadjusted model | | 0.42 |
| Adjusted model | | 0.41 |
| **Parkinson’s disease** | | |
| *Mood disorders* |  | |
| Unadjusted model | 0.44 | |
| Adjusted Model | 0.76 | |
| *Anxiety disorders* |  | |
| Unadjusted model | 0.05 | |
| Adjusted model | 0.21 | |
| *Any mental health disorder* |  | |
| Unadjusted model | 0.09 | |
| Adjusted model | 0.48 | |

**Supplementary Table 5.** Schoenfeld global test *P* values.
